# Digitally managed larviciding as a cost-effective intervention for urban malaria: Operational lessons from a pilot in São Tomé and Príncipe guided by the Zzapp system

**DOI:** 10.1101/2022.09.18.22280065

**Authors:** Arbel Vigodny, Michael Ben Aharon, Alexandra Wharton-Smith, Yonatan Fialkoff, Arnon Houri Yafin, Fernando Bragança, Flavio Soares Da Graça, Dani Gluck, João Alcântara Viegas D’Abreu, Herodes Rompão

## Abstract

**Background:** Once a mainstay of malaria elimination operations, larval source management—the treatment of mosquito breeding habitats–has been marginalized in Africa, due to insufficient effectiveness. However, the development of new technologies, and mosquitoes’ growing resistance to insecticides used in bed nets and house spraying raise renewed interest in this method.

**Methods:** A digitally managed larviciding operation in three of the seven districts of São Tomé and Príncipe (STP) was launched by the Ministry of Health and ZzappMalaria LTD, guided by the Zzapp map-and-GPS mobile application and dashboard. During the operation, quality assurance procedures and field management methods were developed and implemented.

**Findings:** 12,788 water bodies were located and treated a total of 128,864 times. The reduction impact on mosquito population and on malaria cases was 74·90% and 52·65%, respectively. The overall cost per person protected (PPP) was US$0·78 and US$0·41 PPP in the urban areas. Various cost and effectiveness drivers were identified.

**Interpretation:** Digitally managed larviciding can yield highly cost-effective results, especially in urban areas. Digital tools facilitate standardization of operations, implementation of quality assurance procedures, and monitoring of fieldworkers’ performance. Digitally generated spatial data also have the potential to assist integrated vector management operations. A randomized controlled trial with a larger sample is needed to further substantiate findings.

**Funding:** The operation was funded by ZzappMalaria LTD and the STP Ministry of Health (MOH).

## Introduction

Targeting water bodies in which mosquitoes breed—a practice known as larval source management (LSM)—was the mainstay of many malaria control operations in the 1930s and 1940s, often resulting in complete elimination of local malaria transmission.^1^ However, attempts to introduce LSM, and specifically larviciding, to Sub-Saharan Africa were often met with operational difficulties that led to limited coverage and an insufficient impact on reducing mosquito populations. As a result, the current WHO guidelines recommend LSM as a supplementary intervention alongside long-lasting insecticidal nets (LLINs) and indoor residual spraying (IRS), and only in areas where water bodies are “few, fixed, and findable.”^2^

Nevertheless, several factors contribute to a renewed interest in larviciding. First, this method helps mitigate two of the main challenges faced by LLINs and IRS—outdoor biting behavior and insecticide resistance. This, because it affects mosquitoes at their larval stages through biological agents or physical mechanisms to which they are not expected to develop resistance.^3^ In addition, larviciding is potentially highly cost-effective in urban settings,^4^ making it an attractive solution in light of the growing rate of urbanization in sub-Saharan Africa^5^ and of the spread in Africa of the invasive species Anopheles stephensi, which thrive in cities.^6^ Finally, new technologies, e.g., drones and artificial intelligence, can facilitate easier detection and treatment of water bodies.^7^ Similarly, digital tools can promote data-based and data-driven interventions and improve the operational and managerial aspects of large-scale larviciding operations. This paper reports the results of such an operation conducted in São Tomé and Príncipe (STP) by ZzappMalaria LTD and the STP MOH.

## Methods

The Democratic Republic of São Tomé and Príncipe (STP) is an island country in the Gulf of Guinea that consists of two main islands, São Tomé and Príncipe. As of 2021, STP has an estimated population of 228,000,^8^ more than 95% of whom live on the Island of São Tomé. This 854-km^2^ island contains various climatic regions and has a prolonged rainy season that begins in September and lasts through May. The reported number of malaria cases in STP in 2020 was 1933, with an incidence of 8·7 cases per 1000.^9^ The larviciding pilot was performed in three districts: Água Grande, Mé-Zóchi, and Lobata, with a combined area of 243·5km^2^ (28% of the island of São Tomé’s area), and an estimated population of 166,500 people (44% of the country’s population). The districts of Cantagalo, Lembá and Caué, as well as the autonomous island of Príncipe (with a total estimated population of 61,500 people), were not included in the intervention and were therefore used as a control (Fig. 1). While not part of the reported digitized-larviciding intervention, ongoing vector control activities – including IRS, LLINs, drug distribution and community-based larviciding — continued to take place in all the districts.

**Fig. 1:**
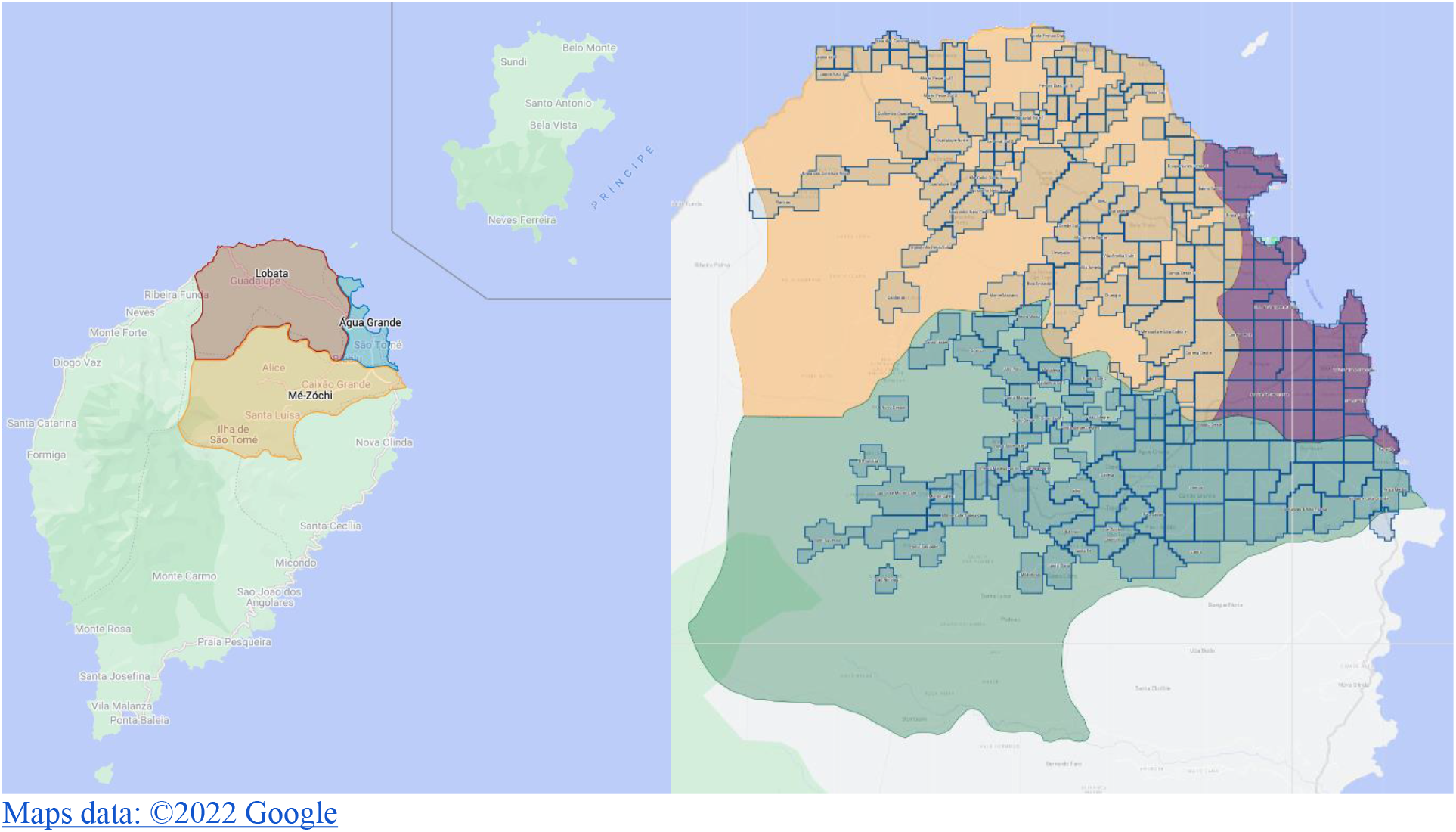
<u>Left</u> – Map of São Tomé and Príncipe, with the intervention districts Água Grande, Lobata and Mé-Zóchi highlighted. <u>Right</u> – Within the three districts, the Zzapp system marked the populated areas and divided them into 255 operational units, with a total area of 125·41 km^2^.

The larviciding operation was guided by the Zzapp system, which comprises an AI-based planning tool that optimizes interventions for any given location, a Mapbox-powered GPS-based mobile app that guides workers in the implementation of the optimized strategies (Fig. 2), and an online dashboard for monitoring operations in real time (Figs. 3, 4). The operation consisted of two phases: a mapping phase in which fieldworkers searched for water bodies, and a treatment phase in which water bodies were treated with larvicide on a weekly basis. The larvicide materials used were VectoBac® G (granules) applied by hand and VectoBac® WDG (water dispersible granules) applied as an aqueous solution. Both products contain the Bacillus thuringiensis var. israelensis bacteria (Bti), which produces toxins targeting a specific protein in the digestive tract of mosquito and black fly larvae, without any harmful effects on other insects and vertebrates.

**Fig. 2:**
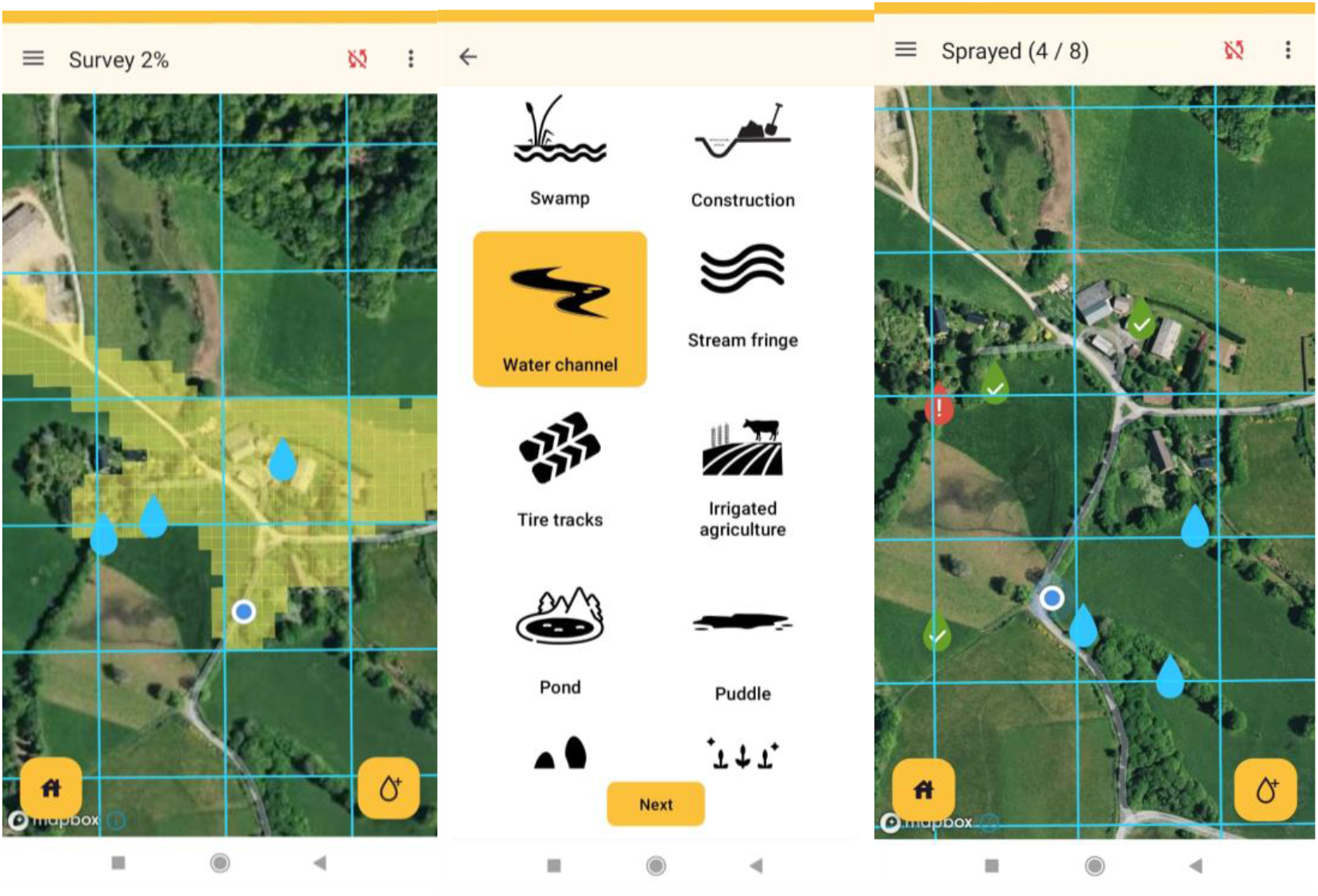
Screenshots from the Zzapp mobile application. Left: Map view during mapping activity showing areas previously visited by the fieldworker highlighted in yellow. The blue circle indicates the current location of the fieldworker, and blue droplet icons indicate water bodies previously reported. Center: Sample questions from the questionnaire completed by fieldworkers for every water body reported. Right: Map view during treatment activity showing droplet icons corresponding to water bodies, color coded by status (green: treated; red: indication of problem preventing treatment; blue: untreated).

**Fig. 3:**
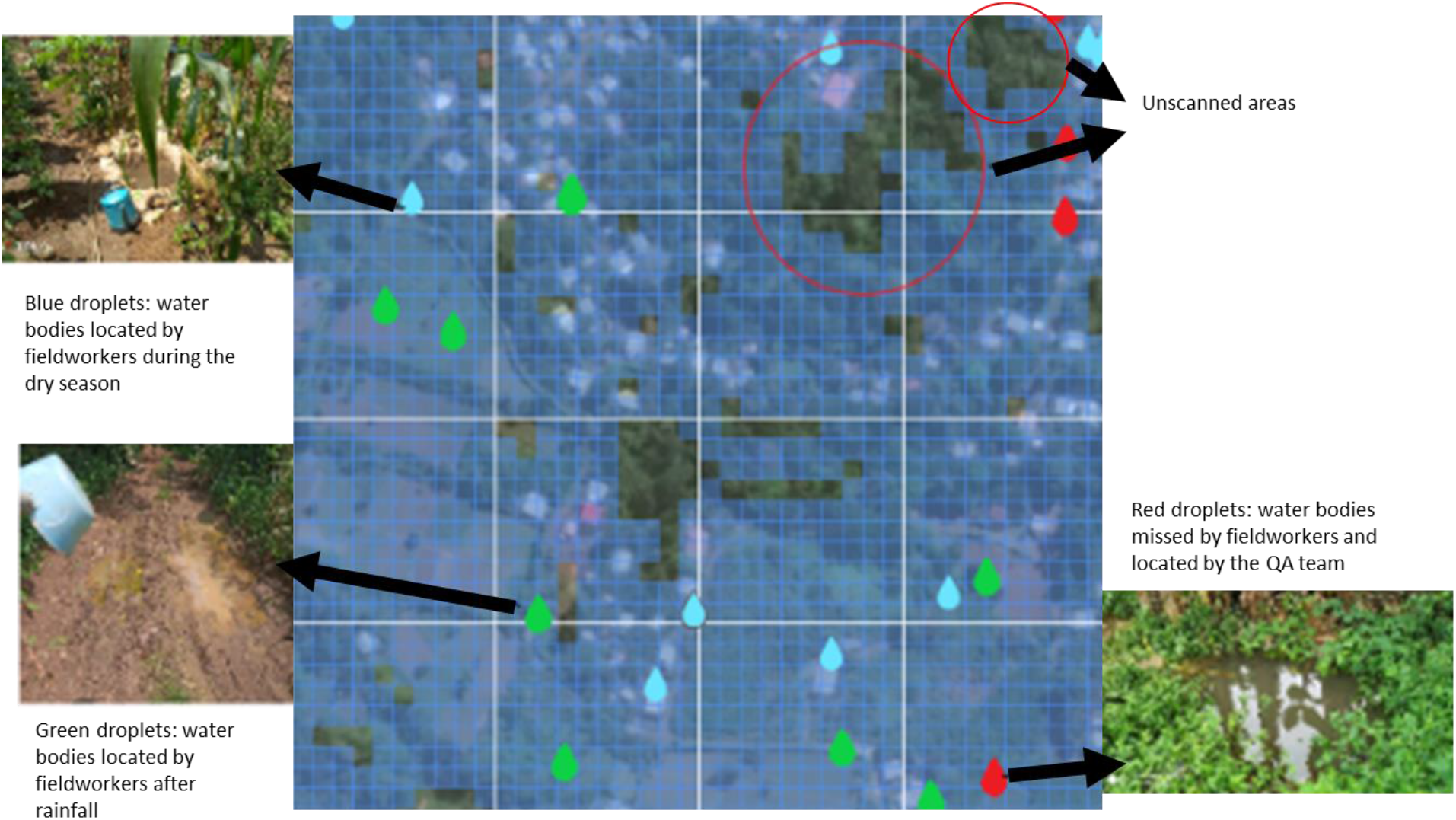
Coverage during the mapping phase. Blue squares indicate areas that were surveyed by fieldworkers (edited from a dashboard screen presenting the village Blublu, Mé-Zóchi district, January 28th, 2022).

**Fig. 4:**
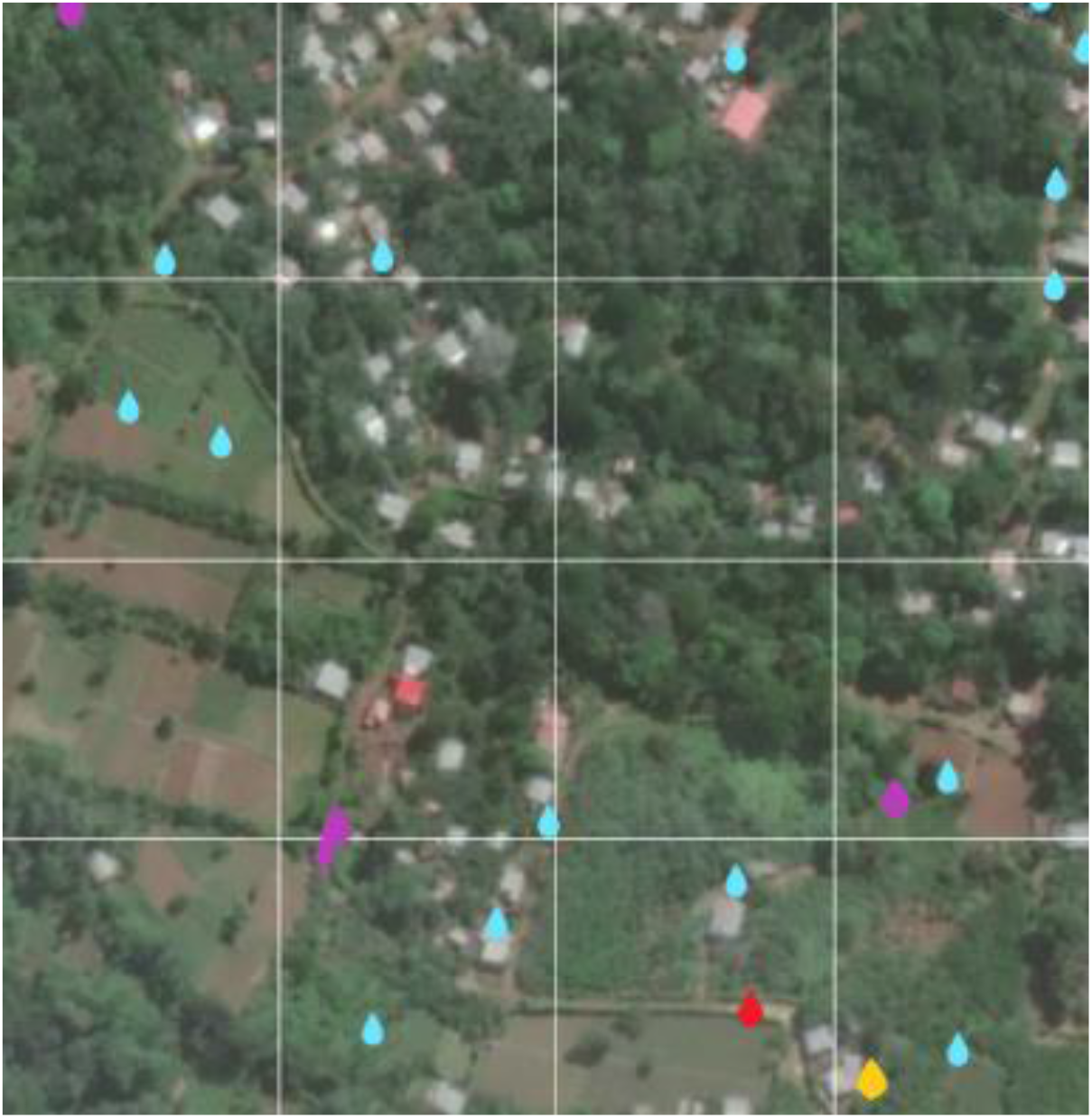
Coverage during the treatment phase. Blue droplets: treated water bodies. Purple droplets: water bodies located during the mapping phase but reported during the treatment phase as nonexistent (e.g., dried out). Orange droplets: water bodies skipped by the fieldworker. Red droplets: water bodies reported as treated but then found positive by the QA team (edited from a dashboard screen presenting the village Blublu, Mé-Zóchi district, January 28th, 2022).

Prior to the implementation stage, fieldworkers underwent a three-day training course, which included an overview of malaria transmission and the life cycle of the anopheles mosquito; the objectives of the operation; guidance on the use of the mobile app in the execution of mapping and application of larviciding of water bodies (including large water bodies); personal safety; field practice; and a practical test. A designated team was trained by an entomologist from the MOH to sample water bodies for mosquito larvae and pupae. Sampling was performed at the beginning of the operation (i.e., prior to treatment of any water bodies) to determine the baseline positivity rate and continued biweekly throughout the operation in the same villages sampled at the baseline. This team repeatedly sampled the same villages in which 150 water bodies testing positive for Anopheles larvae were identified prior to larvicide application (50 positive water bodies per district) in order to monitor the change in positivity over time.

Another group of fieldworkers who were also trained to sample water bodies, served as a quality assurance (QA) team. Their goal was to compliment the Zzapp system in ensuring that the entire area was scanned; that within this area all the water bodies were located; that all water bodies were treated properly (i.e., with the right amount of larvicide at the right frequency); and that all water bodies that appeared in the aftermath of rain were detected. Quality assurance was reached by rescanning certain areas (either by the QA team or by regular fieldworkers) and by sampling, for each fieldworker, a few treated water bodies in order to verify the proper application of larvicide.

Towards the end of the operation, after realizing that the preset milestones for progress were not met, the system was utilized to produce weekly reports to evaluate fieldworkers’ progress with regards to working hours, number of areas assigned for scanning, level of scanning coverage within the assigned areas, and the number of water bodies that were missed or treated insufficiently. In addition, focus group discussions, in-depth interviews, field visits, and informal discussions were carried out, to better understand fieldworkers’ expectations and implementation challenges. As a result, employment structure was rearranged, new agreements specifying working hours and tasks were signed, a bonus system that awarded cash handouts to outstanding workers was established, and workers were provided with daily lunches. These changes correlated to an increase in productivity of 26%.

The operation was piloted to test the Zzapp system in preparation for a nationwide operation in STP and was not designed as a randomized controlled trial. Its effects were measured according to two entomological and one epidemiological criteria: effect on larvae and pupae and adult mosquitoes, and effect on malaria cases. Effect on larvae and pupae was measured through sampling of water bodies performed by fieldworkers trained by an entomologist from the MOH. In each sampling event, five scoops of water were taken from the water body and the larvae and pupae were counted for each scoop, based on their stage of development: Anopheles 1st-2nd instar larvae, Anopheles 3rd-4th instar larvae, Culex/Aedes 1st-2nd instar larvae, Culex/Aedes 3rd-4th instar larvae, and pupae (all species). Sampling was performed at the beginning of the operation (i.e., prior to treatment of any water bodies) to determine the baseline positivity rate, and continued biweekly throughout the operation in the same villages sampled at the baseline. Additionally, the QA team sampled a few water bodies treated by each fieldworker, to verify proper treatment.

The intervention’s effect on the adult mosquito population was measured through routine entomological sampling conducted biweekly by the STP Ministry of Health, using both CDC light traps and human landing collections (HLC). Samples were collected in two locations in each of the country’s seven districts, twice a week, both indoors and outdoors. For each collection point, the ratio between the after and before is an estimator of mosquito population increase in that community. The median of the ratios in all the intervention communities is an estimate of the increase in the entire intervention area. The increase in the control is estimated in the same way. The ratio between the increase in the intervention area and the increase in the control area is the estimate of the intervention impact.

The interventions’ impact on malaria cases relies on official malaria case data, which in STP is collected by the MOH routinely, using weekly reports from health facilities that attribute each malaria case to a location. Given the small number of districts in each group, no significance testing was performed, and no confidence interval was extracted. Rather, the mean number of malaria cases per 10,000 people per week in the entire intervention area and the entire control area were calculated, both in the before-intervention period (weeks 1-49 of 2021) and in the after-intervention period (weeks 1-19 of 2022).

## Findings

### Operational findings

The total area visited by fieldworkers during the ground survey was 90·8 km^2^ (Fig. 5). A total of 12,788 water bodies were reported on the system. These water bodies were treated a total of 128,864 times and sampled 31,353 times. A total of 28,250 “issues” regarding them (e.g., disappearance or lack of accessibility) was reported on the system.

**Fig. 5:**
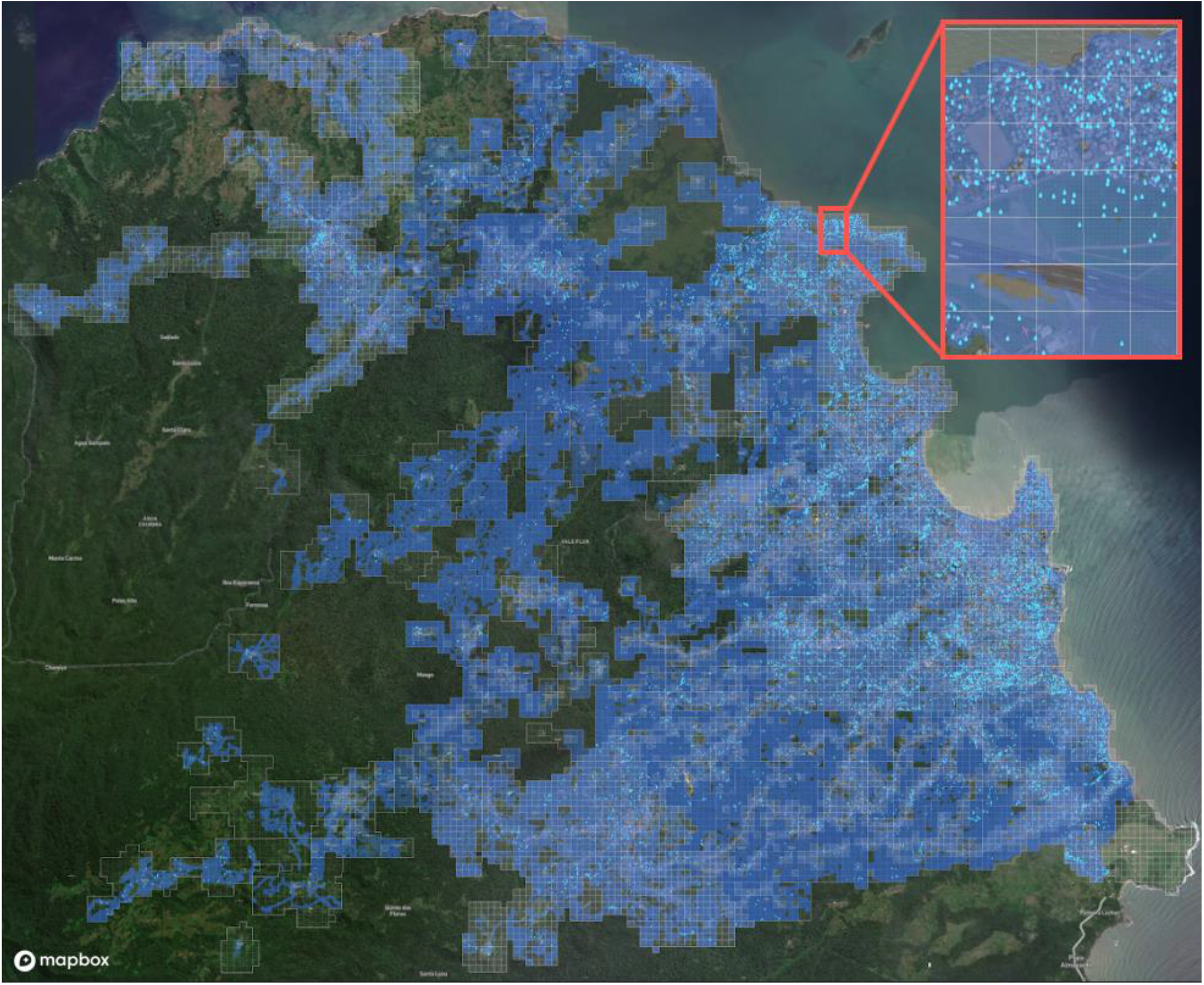
Coverage obtained in mapping activities. The white polygons indicate the area identified for the larviciding intervention. Areas visited by fieldworkers during the mapping stage are highlighted in blue (at a resolution of 10 m^2^). Blue droplet icons mark water bodies reported by fieldworkers.

### Impact on mosquito larvae and pupae

Overall, 31,353 water body samples were collected throughout the operation, showing a decrease of 61·64% in the Anopheles larvae positivity rate during the treatment phase (from 19·42% before the first treatment to 7·44% after 1/12/2022) and a reduction of 81·84% in the pupa positivity rate (from 9·24% before the first treatment to 1·67% after 1/12/2022). Despite this trend, some water bodies remained positive even after the treatment phase, either because they were treated improperly, skipped during the treatment phase, or appeared after the mapping phase (Fig. 6).

**Fig. 6:**
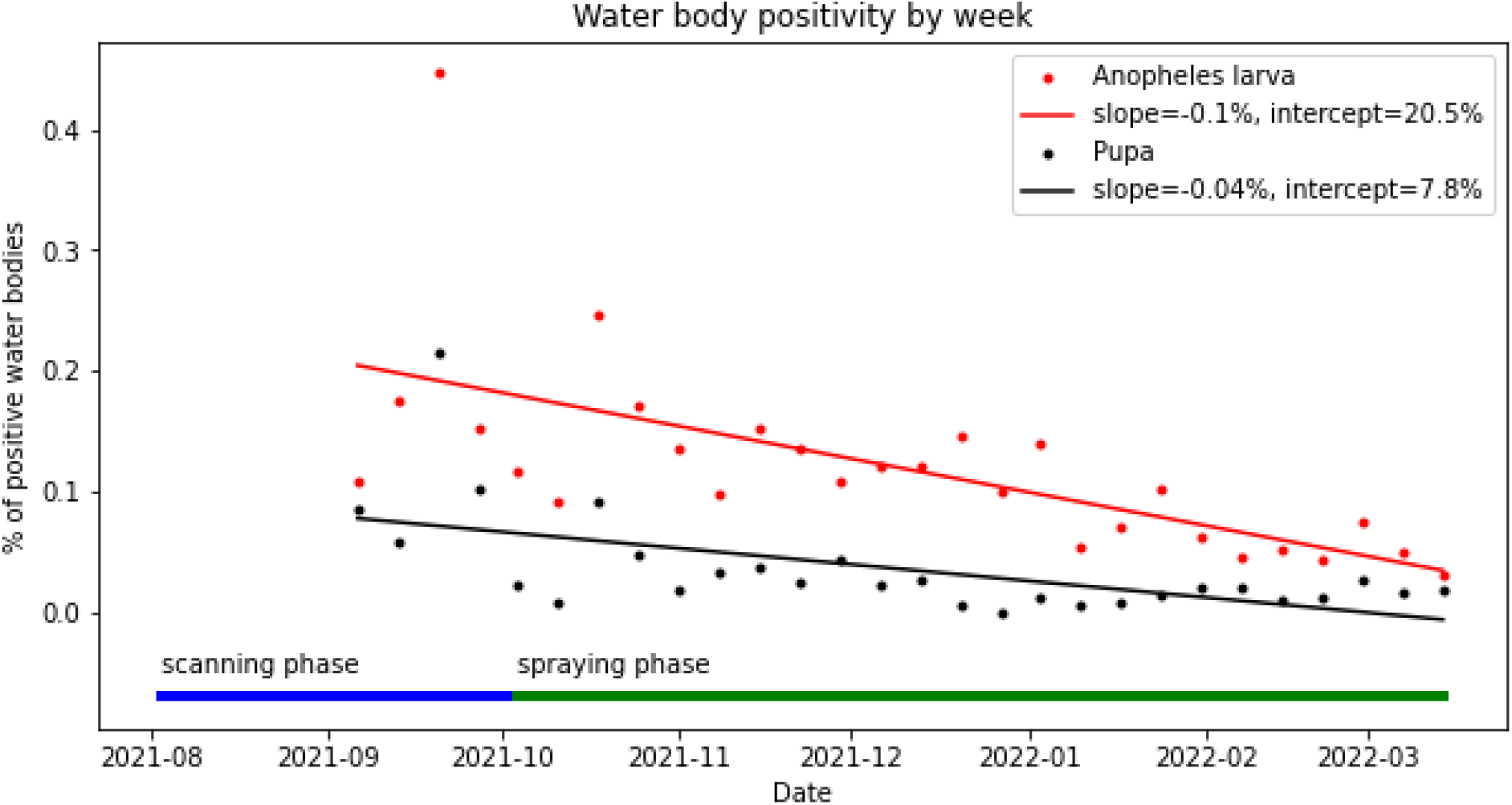
Larva and pupa positivity over the course of the operation.

Table 1 presents the positivity of water bodies before and during the intervention. Water bodies associated with construction were found to have high correlation with pupae positivity. The drivers of mosquito emergence in the area (defined as the multiplication of number of water bodies and positivity rate) were puddles and channels. Tables 2 and 3 present the correlation of certain water body characteristics to pupa positivity before and after treatment. Interestingly, although water pollution was negatively correlated to pupa positivity before treatment, it is positively correlated to pupa positivity after treatment. This may indicate that fieldworkers were not sufficiently trained to treat polluted water bodies with an added amount of Bti, as recommended by the manufacturer.^10^

**Table 1.**
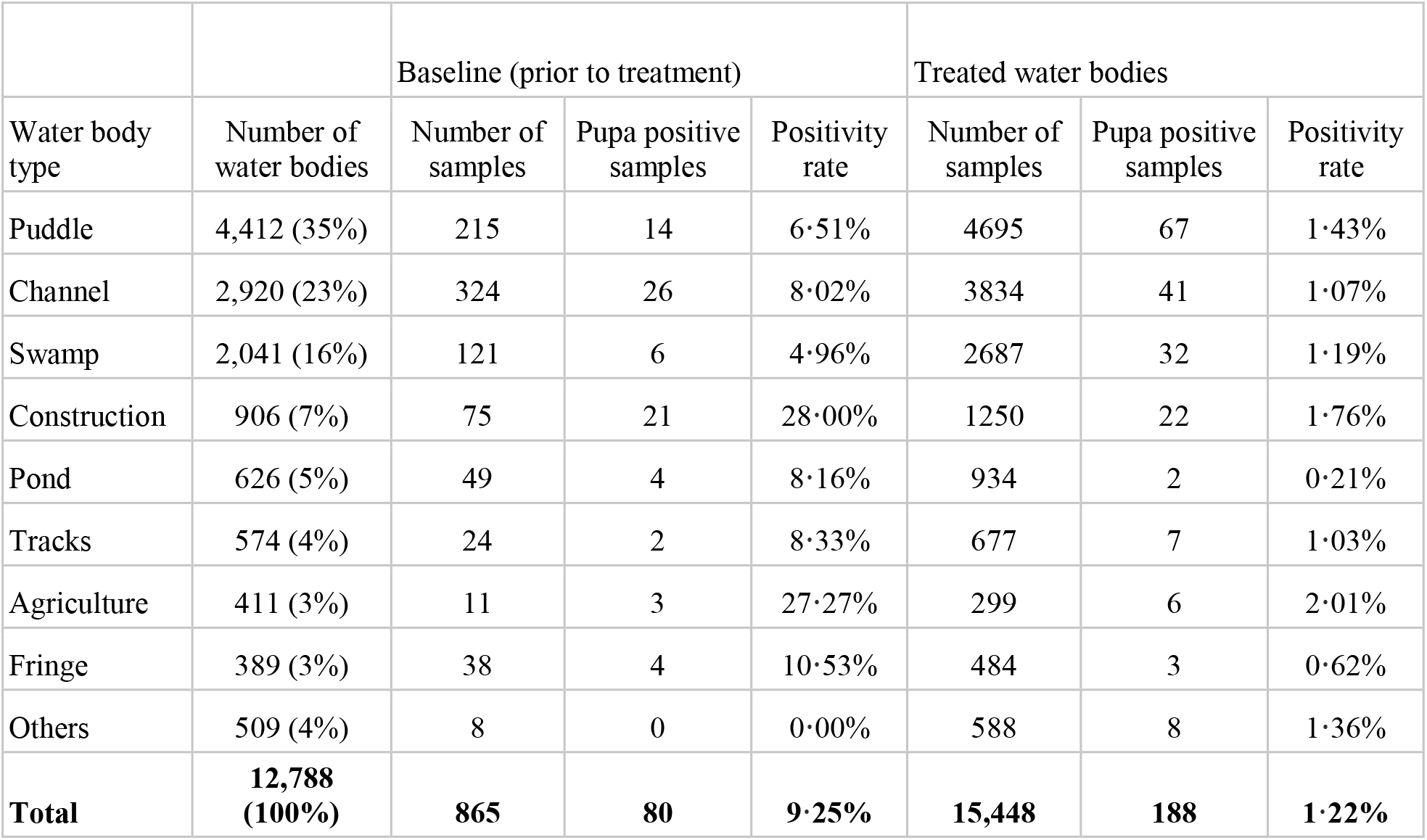
Distribution of water bodies according to type.

**Table 2.** Logistic regression of characteristics of water bodies and positivity before treatment.

|  |  |  |  |  |
| --- | --- | --- | --- | --- |
| Dep. Variable: | Pupa positivity | No. Observations: | 865 |  |
| Model: | Logit | Df Residuals: | 858 |  |
| Method: | MLE | Df Model: | 6 |  |
| Time: | 22:43:10 | Log-Likelihood: | -251.12 |  |
|  | coef | std err | Z | P |
| const | -1.7625 | 0.249 | -7.09 | 0 |
| isPolluted | -0.3908 | 0.243 | -1.61 | 0.108 |
| isShaded | -0.5619 | 0.255 | -2.21 | 0.027 |
| isVegetation | -0.3226 | 0.267 | -1.21 | 0.227 |
| isTemporary | -0.7578 | 0.283 | -2.68 | 0.007 |
| depth (cm) | 0.0150 | 0.005 | 3.12 | 0.002 |
| area (meter) | -0.0398 | 0.03 | -1.34 | 0.18 |

**Table 3.** Logistic regression of water bodies characteristics and positivity after treatment.

|  |  |  |  |  |
| --- | --- | --- | --- | --- |
| **Dep. Variable:** | Pupa positivity | **No. Observations:** | 23,607 |  |
| **Model:** | Logit | **Df Residuals:** | 23,600 |  |
| **Method:** | MLE | **Df Model:** | 6 |  |
|  | **coef** | **std err** | **z** | **p** |
| **const** | -4.309 | 0.128 | -33.71 | 0 |
| **isPolluted** | 0.2635 | 0.114 | 2.30 | 0.021 |
| **isShaded** | -0.1307 | 0.111 | -1.18 | 0.239 |
| **isVegetation** | -0.1204 | 0.116 | -1.04 | 0.297 |
| **isTemporary** | -0.0053 | 0.115 | -0.05 | 0.963 |
| **depth (cm)** | 0.0026 | 0.002 | 1.10 | 0.273 |
| **area (meter)** | -0.0042 | 0.014 | -0.30 | 0.763 |

An additional factor accounting for positivity of water bodies may have been insufficient frequency of treatment. Fig. 7 shows the correlation between positivity of water bodies and the number of days since the last treatment. In this pilot, the minimum time between treatment events was set as five days, the target interval as seven days, and the maximum interval as 14 days, after which the system would alert the operation administrator through the dashboard. In the operation, the average interval between visits was 10·8 days, which may explain the positivity of some water bodies.

**Fig. 7:**
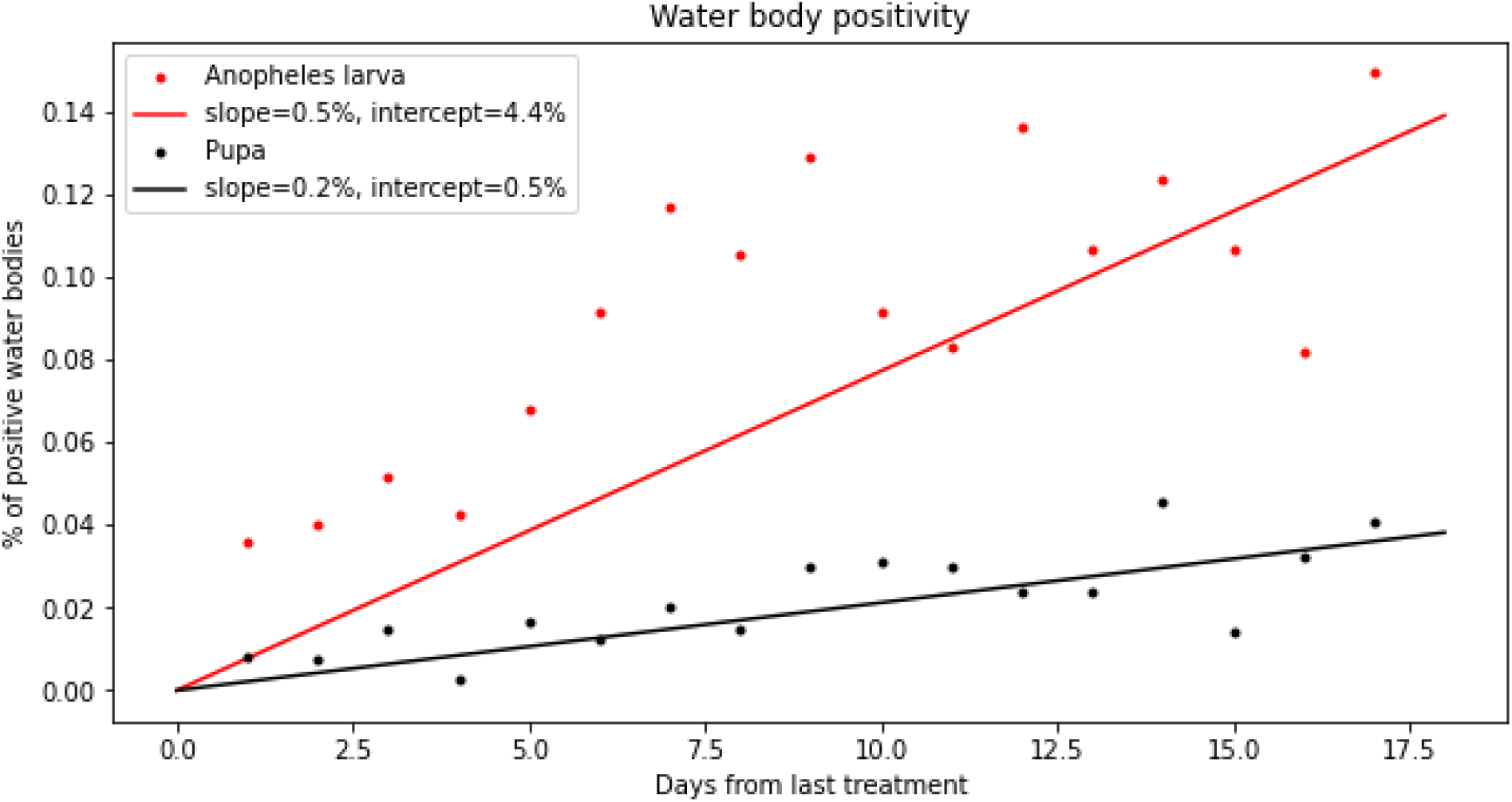
Sampling of water bodies by the QA team. Note that the baseline positivity (before spraying) for larva is 19·3% and for pupae is 9·2%. Even after 17 days, the treatment has some impact on water body positivity for Anopheles mosquitoes.

### Impact on adult mosquitoes

Indoor collections, both by HLC and by CDC light traps, produced very low Anopheles counts compared with outdoor collections. The monthly averages of Anopheles mosquitoes collected for all collection points combined was 3·3 (indoors) vs. 55·6 (outdoors) for light traps and 10·6 (indoors) vs. 248·8 (outdoors) for HLC. Since outdoor light traps gave relatively low numbers, with 5 out of the 14 points collecting 0 mosquitoes during both the “before” and “after” periods, an after-before ration comparison was impossible (outdoor HLC collection points did not have this problem). Results are summarized in Table 4.

**Table 4:** Total Anopheles outdoor HLC, before and after intervention.

| Community | Total outdoor *Anopheles* HLC, before intervention | Total outdoor *Anopheles* HLC, after intervention | Ratio after/before |
| --- | --- | --- | --- |
| Intervention communities |  |  |  |
| Praia Gamboa | 385 | 35 | 0·091 |
| Madre De Deus | 25 | 1 | 0·040 |
| Cidade De Cruzeiro | 9 | 0 | 0·000 |
| Cidade De Praia Melão | 128 | 3 | 0·023 |
| Micolo | 209 | 85 | 0·407 |
| Conde | 178 | 48 | 0·270 |
| Median |  |  | **0·065** |
| control communities |  |  |  |
| Cidade Das Neves | 138 | 98 | 0·710 |
| Ponta Figo | 47 | 1 | 0·021 |
| Ribeira Afonso | 412 | 36 | 0·087 |
| Zandrigo | 84 | 152 | 1·810 |
| Porto Real (Príncipe) | 201 | 65 | 0·323 |
| Rua Dos Trabalhadores (Príncipe) | 208 | 54 | 0·260 |
| Angolares | 3 | 0 | 0·000 |
| Emolve | 305 | 79 | 0·259 |
| Median |  |  | 0·259 |

The medians for the after/before ratios of the intervention and control groups were 0·065 and 0·259, respectively, giving a relative change of -74·90%, with a one-sided 75% confidence interval of (-100%, -30%), estimated using bootstrapping. Note that a 75% confidence interval was proposed for clinical pilot studies.^11,12^ This result requires confirmation in a randomized control trial with a sample size powered to detect impact of at least 30%. It is important to note, however, that this result remains robust in other analysis methods—using means instead of medians, including indoor collections using light traps instead of HLC.

### Impact on malaria cases

The mean number of malaria cases per 10,000 people per week in all the intervention districts combined was 2·86 in the before-intervention period and 4·82 in the after-intervention period, giving an after/before ratio of 1·69. The mean number of malaria cases per 10,000 people per week in all the control districts combined was 0·86 in the before-intervention period and 3·08 in the after-intervention period, giving a ratio of 3·57. Thus, the after/before ratio in the intervention districts is 52·65% smaller than the control districts.

### Cost

The total cost of the operation, including a two-month mapping stage and a 5·5-month larviciding stage, was US $129,874. Cost categories are detailed below (Table 5). The main cost drivers were labor, transportation and larvicide material (similar to Worrall et al.).^13^ The overall cost of the operation per person protected (PPP) was US$ 0·78. Cost varied significantly with population density.

**Table 5:**
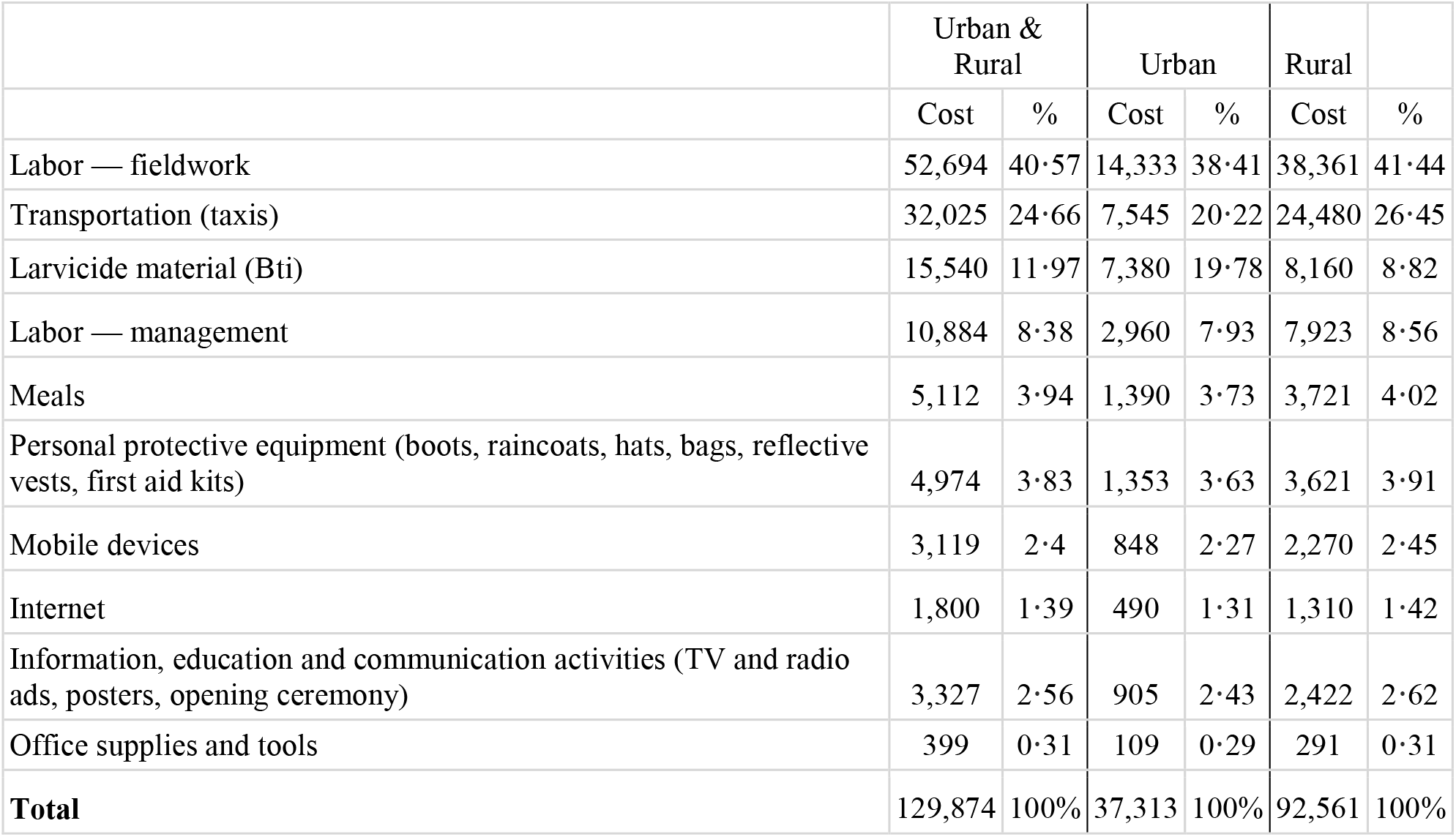
Cost categories divided between urban and rural areas (US$)

|  | Urban & Rural |  | Urban |  | Rural |  |
| --- | --- | --- | --- | --- | --- | --- |
|  | Cost | % | Cost | % | Cost | % |
| Labor — fieldwork | 52,694 | 40.57 | 14,333 | 38.41 | 38,361 | 41.44 |
| Transportation (taxis) | 32,025 | 24.66 | 7,545 | 20.22 | 24,480 | 26.45 |
| Larvicide material (Bti) | 15,540 | 11.97 | 7,380 | 19.78 | 8,160 | 8.82 |
| Labor — management | 10,884 | 8.38 | 2,960 | 7.93 | 7,923 | 8.56 |
| Meals | 5,112 | 3.94 | 1,390 | 3.73 | 3,721 | 4.02 |
| Personal protective equipment (boots, raincoats, hats, bags, reflective vests, first aid kits) | 4,974 | 3.83 | 1,353 | 3.63 | 3,621 | 3.91 |
| Mobile devices | 3,119 | 2.4 | 848 | 2.27 | 2,270 | 2.45 |
| Internet | 1,800 | 1.39 | 490 | 1.31 | 1,310 | 1.42 |
| Information, education and communication activities (TV and radio ads, posters, opening ceremony) | 3,327 | 2.56 | 905 | 2.43 | 2,422 | 2.62 |
| Office supplies and tools | 399 | 0.31 | 109 | 0.29 | 291 | 0.31 |
| **Total** | **129,874** | **100%** | **37,313** | **100%** | **92,561** | **100%** |

Of the total scanned area, 12·87% (16·15 km^2^) was urban (>1,500 structures per square km, based on Open Buildings dataset),^14^ in which an estimated 56·31% of the total intervention population (93,762 people) live. For higher resolution of the correlation between population density and cost PPP, see Fig 7. According to data from the mobile application, 27·2% of workdays, and 47·5% of treatment events, took place in these urban localities. The cost in urban areas was an estimated US$ 37,313, and US$ 0·41 PPP, and the cost in rural areas was an estimated US$ 92,561, and US$ 1·23 PPP (see Fig. 8). Appendix 1 presents an elaborate calculation of cost and the cost saving that could be achieved by using operation-owned cars instead of taxis.

**Fig. 8:**
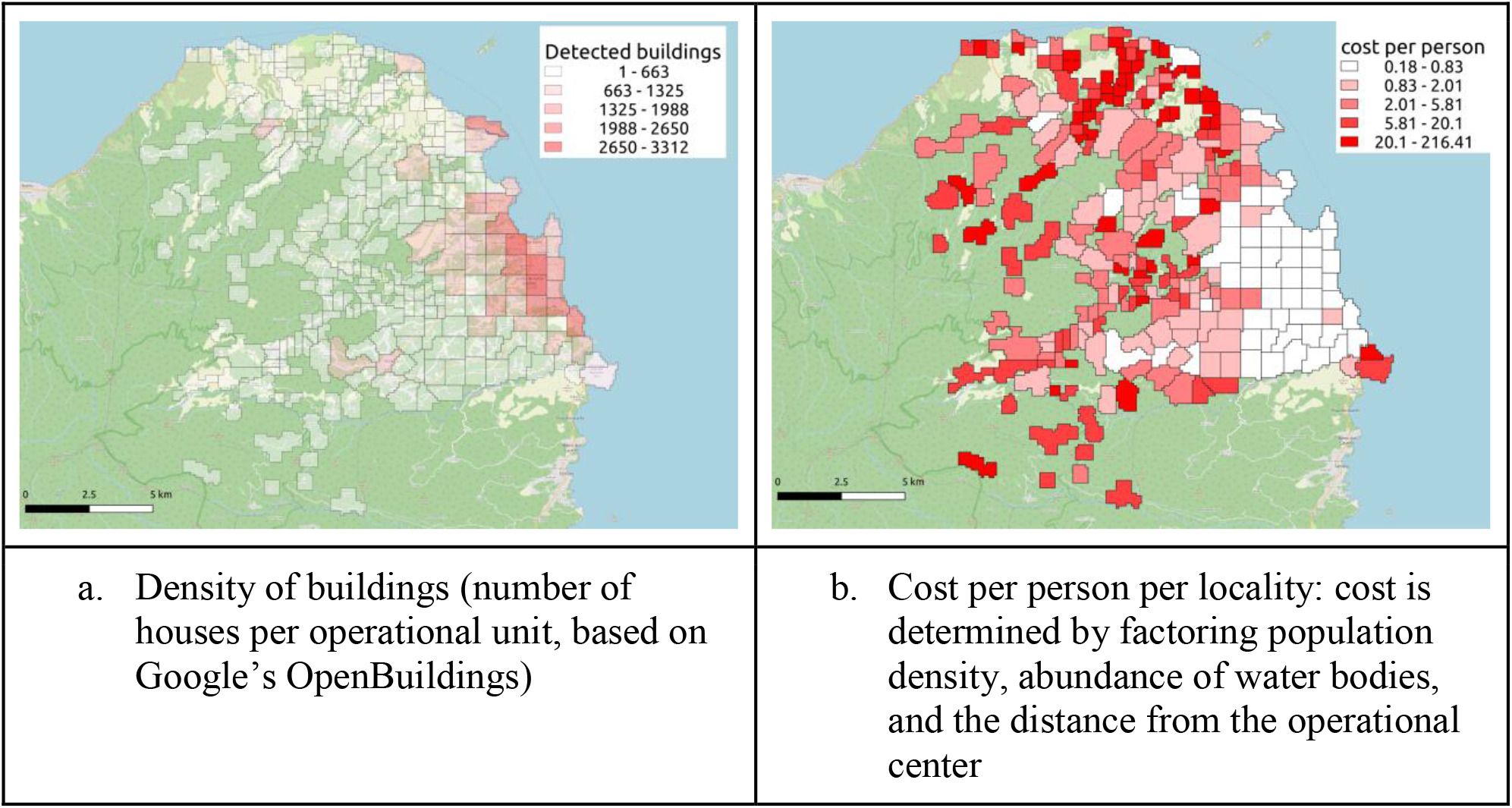
Comparison of a) population density (based on number of buildings per km^2^) and b) cost (labor, transportation, and Bti) PPP per locality.

## Discussion

The findings of the operation reported here indicate the high cost-effectiveness of digitally managed larviciding (DML) operations, especially in urban and semi-urban areas. It is important to note that there is a need to confirm these findings by randomized controlled trials conducted in varied settings and with larger sample sizes. If in fact confirmed, the effectiveness that was achieved—52·65% reduction of malaria cases—is comparable to that reported in studies measuring the effectiveness of LLINs (45%^15^) and IRS (18%)^16^; the cost, however, was significantly lower: US$ 0·359 per person protected per 6 months compared with US$ 0·695^17^ and US$ 6·19^18^ respectively.

Moreover, as Appendix 1 elaborates, there is substantial room for improvement of both cost and effectiveness of DML. Crucially, digitization enables effective monitoring of the operation, making operations more standardized and replicable.

At the same time, new tools can increase the cost-effectiveness of DML, making it a suitable solution for a greater number of areas. For example, drones and detection of water bodies from satellite imagery using AI can be used to optimize the detection and treatment of water bodies,^19,20^ and analysis of weather conditions and patterns may help to choose the best timing for interventions.

Such improvements, along with spatial modeling and data about water body location, can also be optimize the use of other methods, e.g. recommending the houses to be treated with IRS or where to place attractive targeted sugar baits (ATSBs).^21^ Thus, digitization may be the enabler of long-sought, but seldom implemented, integrated vector management (IVM) operations.^22^

The key for all the above is efficient monitoring mechanisms that provide reliable and granular data in real time. During operations, monitoring enables tracking progress, evaluating workers’ performance, flagging areas deserving extra attention, and expanding the use of successful interventions. Monitoring also facilitates tracking of expenses, thereby both helping to reduce operational costs and providing clear, detailed and precise accountability reports to stakeholders. Finally, close monitoring strengthens the robustness of results, yielding reliable data-based insights and recommendations for future research and interventions. Digitization facilitates the pooling and analysis of various malaria data and on multiple levels – from the location of a water body in a village to the average distance to hospitals in a given district, from transportation costs to community acceptance. Aggregating this information into a single, spatial-based platform can significantly improve vector control operations, to the extent of reproducing the results of historical LSM operations: nationwide elimination.

## Data Availability

All data produced in the present study are available upon reasonable request to the authors

## Appendix 1. Cost

Following Worrall’s ingredients approach,^13^ the overall financial cost of the operation was calculated by identifying each activity involved in the operation from the perspective of the intervention provider, and summing their costs. Data sources used for the cost estimation include expense reports maintained by the STP MOH, National Center for Endemic Diseases (Centro Nacional de Endemias; CNE) and ZzappMalaria, fieldworker attendance logs, transportation usage logs, and mobile device inventory logs. Access to the mobile application, training for project managers, and ongoing support for the Zzapp system were provided by ZzappMalaria at no cost, and thus not included in the estimate. Office space was available at the CNE prior to commencement of the operation, and was provided at no cost, and is thus excluded from the estimate. Research costs that would not normally be required for implementation of the larviciding intervention (e.g., increased entomological monitoring or field visits by the ZzappMalaria team) were also excluded. Expenses were converted to USD based on the exchange rate at the date of payment or using the average exchange rate in the relevant period.

Capital costs were calculated based on the predicted duration of use of budget items expected to last longer than the duration of the operation (7·5 months in total). The only capital cost was for the purchase of mobile devices for use by the fieldworkers. PPE and other equipment costs were not considered capital costs due to their high replacement rates.

Costs were further divided into those associated with operating in urban areas vs. rural areas (Table 5 in the main paper). Each locality was defined as urban or rural based on the number of structures per km^2^ (>1,500), which was calculated using Google’s Open Buildings dataset.^23^ Data from the Zzapp mobile application were then used to determine the number of workdays spent in urban vs. rural localities, out of the total workdays registered. Labor costs and derivative costs, including management, equipment and supplies, internet usage, and IEC activities were divided into urban or rural based on the workdays ratio. Larvicide usage was divided based on data from the Zzapp application, which records individual treatment events by water body location and size. Surprisingly, water bodies in urban and rural area had similar size distribution; furthermore, the number of water bodies per km^2^ was higher in the urban areas. Transportation costs were divided based on daily transportation logs.

The financial cost PPP was calculated by dividing the total cost of the operation by the estimated population size in the three intervention districts, obtained from official government statistics. The urban and rural population sizes were estimated by determining the total number of structures in each category, as described above, and multiplying the resulting ratio by the total intervention population (assuming similar occupancy per structure across the intervention districts).

### Spatial mapping of costs

The cost of larviciding varies according to the prevalence of water bodies (which affects the amount of labor and Bti); population density within villages (which affects the amount of area to be scanned); and the distance between the villages and the operational center (which affects the consumption of fuel and fieldworkers’ transportation time). Table 1 compares various parameters of rural vs. urban localities in the intervention area in the operation in STP.

**Table 1.** Rural vs. Urban localities in the intervention area.

|  | Rural |  | Urban |  |
| --- | --- | --- | --- | --- |
|  | # | Of total | # | Of total |
| Localities | 251 | 90·61% | 26 | 9·39% |
| Total area | 122·92 km<sup>2</sup> | 88·39% | 16·15 km<sup>2</sup> | 11·61% |
| Water bodies reported | 7,324 | 57·27% | 5,464 | 42·73% |
| Houses | 37,391 | 44·73% | 46,198 | 55·27% |
| Estimated population | 72,738 | 43·69% | 93,762 | 56·31% |
| Water body treatment events | 67,656 | 52·50% | 61,208 | 47·49% |
| Workdays | 3,594 | 72·83% | 1,341 | 27·17% |

Fig. 8 (in the main paper) presents a map of costing, based on data from the app’s log to factor the labor per village and the distance between villages to the operational center in order to assess the cost per location. As the map shows, the cost varies, with a few “villages” (actually lone houses and the isolated “voice of America” radio station in the southwest) costing more than $200 PPP. It is important to note that while this tool is meant to assist managers in assessing costs and prescribing the appropriate interventions to each area, it is by no means intended to discriminate against any locality or any population. Areas for which larviciding is too expensive must be treated with alternative methods that do not require frequent visits—for example IRS and window screening—and are therefore less costly.

### Options for cost reduction

A significant improvement of cost-effectiveness could be achieved by treating the water bodies during the scanning phase rather than in a separate treatment phase. Doing so, is expected to prolong the impact of the operation with minimal additional cost (although it interferes with establishing an entomological baseline to monitor the operations results).

Due to the limited duration of the pilot, use was made of taxis instead of procuring cars, which significantly increased the cost of transportation. In preparation for expansion of the pilot to cover the entire island of São Tomé, we conducted a cost estimate for transportation using cars purchased for the project. Used vehicles suitable for the project requirements and the terrain on the island were searched online to determine availability and estimate cost. Logs from the mobile app were analyzed to determine the typical distance traveled per day by the teams, the number of cars that would be needed to make the number of trips observed, and the number of kilometers traveled. Gas mileage was estimated based on averages published online for the cars selected. Maintenance and insurance costs were estimated by managers in São Tomé based on their experience with similar cars. Driver salaries were determined based on the rate paid by the MOH for drivers. Assuming a 10-year usable life for the cars, the estimated cost including fuel and salary for drivers was estimated to be between US$ 22,045 per year, and 14,772 for a period of 7·5 months (the duration of the trial)—less than half of what was actuality spent (table 2).

**Table 2:** Estimated transportation costs (US$)

| Capital Costs |  |  |  |  |  |
| --- | --- | --- | --- | --- | --- |
|  | Cost per unit | Units | Usable life (years) | Cost per year | Cost per 7·5 months |
| Vehicles | 23,000 | 3 | 10 | 6,900 | 5,175 |
| Recurrent Costs |  |  |  |  |  |
|  | Cost per unit | Units | Period | Cost per year | Cost per 7·5 months |
| Fuel | 1·3 | 25/day | Day (20/month) | 7,800 | 4,875 |
| Drivers | 7·91 | 3/day | Day (20/month) | 5,695 | 3,559 |
| Maintenance | 350 | 3 | Year | 1,050 | 788 |
| Insurance | 200 | 3 | Year | 600 | 375 |
| Total |  |  |  | 22,045 | 14,772 |

## References

1 Killeen GF, Fillinger U, Kiche I, Gouagna LC, Knols BG. Eradication of Anopheles gambiae from Brazil: lessons for malaria control in Africa? Lancet Infect Dis. 2002; 2(10): 618–27.

2 World Health Organization, Larval Source Management: a supplementary measure for malaria vector control, an operational manual. chrome-extension://efaidnbmnnnibpcajpcglclefindmkaj/http://apps.who.int/iris/bitstream/handle/10665/85379/9789241505604_eng.pdf;jsessionid=E018E2C53CF30F460864EFC577D76BF8?sequence=1.

3 Tetreau G, Stalinski R, David JP, Després L. Monitoring resistance to Bacillus thuringiensis subsp. israelensis in the field by performing bioassays with each Cry toxin separately. Memórias do Instituto Oswaldo Cruz. 2013; 108: 894–900.

4 Maheu-Giroux M, Castro MC. Cost-effectiveness of larviciding for urban malaria control in Tanzania. Malaria journal. 2014; (1): 1–2.

5 The World Bank. Urbanization in Africa: Trends, Promises, and Challenges. The World Bank Group, https://www.worldbank.org/en/events/2015/06/01/urbanization-in-africa-trends-promises-and-challenges. 2015.

6 Sinka ME, Pironon S, Massey NC, Longbottom J, Hemingway J, Moyes CL, Willis KJ. A new malaria vector in Africa: Predicting the expansion range of Anopheles stephensi and identifying the urban populations at risk. Proceedings of the National Academy of Sciences. 2020; 117(40):24900–8.

7 Hardy A, Makame M, Cross D, Majambere S, Msellem M. Using low-cost drones to map malaria vector habitats. Parasites & vectors. 2017; 10(1):1–3.

8 https://databank.worldbank.org/source/population-estimates-and-projections

9 World Health Organization. World malaria report 2021. World Health Organization, 2021.

10 https://www.valentbiosciences.com/publichealth/products/vectobac/

11 Lee EC, Whitehead AL, Jacques RM, Julious SA. The statistical interpretation of pilot trials: should significance thresholds be reconsidered?. BMC medical research methodology. 2014; 14(1): 1–8.

12 Schoenfeld D. Statistical considerations for pilot studies. International Journal of Radiation Oncology· Biology· Physics. 1980; 1;6(3): 371–4. Schoenfeld, D. (1980). “Statistical considerations for pilot studies.” International Journal of Radiation Oncology - Biology - Physics, 6(3), 371–374.

13 Worrall E, Fillinger U. Large-scale use of mosquito larval source management for malaria control in Africa: a cost analysis. Malaria journal. 2011; 10(1): 1–21.

14 https://sites.research.google/open-buildings

15 Pryce J, Richardson M, Lengeler C. Insecticide-treated nets for preventing malaria. Cochrane Database of Systematic Reviews. 2018; (11): 1–66.

16 Pryce J, Medley N, Choi L. Indoor residual spraying for preventing malaria in communities using insecticide-treated nets. Cochrane Database of Systematic Reviews. 2022; (1): 1–83.

17 Based on a division in half of the median economic cost PPP per year of LLINs (US$1·39) and IRS (US$5·70) according to Conteh, Lesong, et al. Conteh L, Shuford K, Agboraw E, Kont M, Kolaczinski J, Patouillard E. Costs and cost-effectiveness of malaria control interventions: a systematic literature review. Value in Health. 2021; 24(8): 1213–22.

18 Johns B, Mignote H. PMI IRS Country Programs: 2019 Comparative Cost Analysis. Rockville, MD. PMI VectorLink Project, Abt Associates Inc. 2020.

19 Mukabana WR, Welter G, Ohr P, Tingitana L, Makame MH, Ali AS, Knols BG. Drones for Area-Wide Larval Source Management of Malaria Mosquitoes. Drones. 2022; 6(7): 180.

20 Talal M, Panthakkan A, Mukhtar H, Mansoor W, Almansoori S, Al Ahmad H. Detection of water-bodies using semantic segmentation. In 2018 International Conference on Signal Processing and Information Security (ICSPIS) 2018; 1–4.

21 White MT, Griffin JT, Churcher TS, Ferguson NM, Basáñez MG, Ghani AC. Modelling the impact of vector control interventions on Anopheles gambiae population dynamics. Parasites & vectors. 2011 Dec; 4(1): 1–4.

22 Beier JC, Keating J, Githure JI, Macdonald MB, Impoinvil DE, Novak RJ. Integrated vector management for malaria control. Malaria journal. 2008; 7(1): 1–0.

23 https://sites.research.google/open-buildings/

